# Unmasking Neuroendocrine Prostate Cancer with a Machine Learning-Driven 7-Gene Stemness Signature that Predicts Progression

**DOI:** 10.1101/2024.09.24.24314303

**Authors:** Agustina Sabater, Pablo Sanchis, Rocio Seniuk, Gaston Pascual, Nicolas Anselmino, Daniel Alonso, Federico Cayol, Elba Vazquez, Marcelo Marti, Javier Cotignola, Ayelen Toro, Estefania Labanca, Juan Bizzotto, Geraldine Gueron

## Abstract

Prostate cancer (PCa) poses a significant global health challenge, particularly due to its progression into aggressive forms like neuroendocrine prostate cancer (NEPC). This study developed and validated a stemness-associated gene signature using advanced machine learning techniques, including Random Forest and Lasso regression, applied to large-scale transcriptomic datasets. The resulting 7-gene signature (*KMT5C, MEN1, TYMS, IRF5, DNMT3B, CDC25B and DPP4*) was validated across independent cohorts and patient-derived xenograft (PDX) models. The signature demonstrated strong prognostic value for progression-free, disease-free, relapse-free, metastasis-free, and overall survival. Importantly, the signature not only identified specific NEPC subtypes, such as large-cell neuroendocrine carcinoma, which is associated with very poor outcomes, but also predicted a poor prognosis for PCa cases that exhibit this molecular signature, even when they were not histopathologically classified as NEPC. This dual prognostic and classifier capability makes the 7-gene signature a robust tool for personalized medicine, providing a valuable resource for predicting disease progression and guiding treatment strategies in PCa management.

## INTRODUCTION

Prostate cancer (PCa) remains one of the most significant health challenges for men globally, with a high incidence and mortality, particularly in advanced stages of the disease [1]. Despite advancements in early detection and treatment, accurately predicting which patients will experience aggressive disease progression remains a major challenge. A critical gap in the management of PCa is the lack of reliable prognostic biomarkers capable of identifying patients at the highest risk of developing more aggressive forms of PCa, such as neuroendocrine prostate cancer (NEPC), a subtype associated with poor prognosis [2,3]. Addressing this gap is essential for improving patient outcomes and guiding effective therapeutic strategies.

To overcome this challenge, we propose the identification of stem-like characteristics within prostate tumors. Cancer stem cells (CSCs) are a subpopulation of cells within tumors that possess the ability to self-renew, differentiate, and drive tumor growth, metastasis, and resistance to conventional therapies [2,4]. These cells have been implicated in the recurrence and progression of PCa, making them critical targets for both prognostic and therapeutic interventions [5]. Moreover, CSCs are believed to contribute to the heterogeneity of PCa, which complicates treatment and highlights the need for more refined biomarkers [6]. However, despite the recognized importance of CSCs, there is still a need for concise and clinically applicable biomarkers, such as transcriptomic signatures, that can reliably point out the presence of stemness traits in prostate tumors and their associated risk of progression.

In addition to their role in driving tumor growth, CSCs are more abundant after the neuroendocrine differentiation of prostate cancer, which results in NEPC [7]. This PCa subtype can arise either de novo or through the transdifferentiation of adenocarcinoma under selective pressures such as androgen deprivation therapy (ADT) [8,9]. This transdifferentiation process, driven by cellular plasticity and epigenetic changes, results in a highly aggressive cancer subtype that is associated with poor outcomes and limited treatment options [10,11]. Identifying biomarkers that can detect early shifts toward a neuroendocrine phenotype is crucial for managing treatment-resistant cases. However, existing NEPC-related gene signatures are often complex, including a large number of genes, limiting their practical use in clinical settings [12,13].

In this study, we aimed to address these challenges by developing a concise and robust stemness-associated gene signature using machine learning techniques. By analyzing large-scale transcriptomics data from multiple cohorts, we identified a 7-gene signature that predicts multiple PCa disease progression events. This signature was rigorously validated across independent datasets and further substantiated using patient-derived xenograft (PDX) models and a NEPC dataset, where we observed that our signature is able to classify samples as NEPC and, particularly, the large cell neuroendocrine carcinoma subtype. Our comprehensive approach provides a novel and clinically applicable tool for patient stratification and treatment personalization, offering new insights into the role of stem-like traits in PCa and their association with neuroendocrine differentiation.

## MATERIALS AND METHODS

### Stemness-associated genes

We gathered 144 stemness-associated genes from PCa literature [6,14–18]. We conducted transcriptomics analyses using publicly available PCa datasets (see below) to study differential gene expression across multiple comparisons, including normal/benign tissues, primary PCa tumors, CRPC tumors and metastatic samples. We performed univariable survival analysis to study the association between gene expression and different endpoints (progression, disease-free time biochemical-relapse, metastasis, and death). We also performed multivariable survival analyses that included clinico-pathological features as covariables.

### Transcriptomics analyses

#### Dataset selection criteria

To study differential gene expression across different PCa datasets, we searched Gene Expression Omnibus (GEO) and the Genomic Data Commons Data Portal to identify eligible datasets that met the following criteria: (1) PCa tissue samples with available transcriptomic and clinico-pathological data; (2) The datasets must have ≥2 different tissue sample types (**Table 1**).

**Table 1:** PCa transcriptomics datasets for differential expression analysis.

| <u>Dataset</u> | <u>Samples</u> |
| --- | --- |
| GSE35988 [19] | Localized PCa (n=59), matched benign prostate tissues (n=28), and metastatic CRPC (n=35). |
| GSE3933 [20,21] | Localized PCa (n=62) and normal prostate (n=41). |
| GSE46602 [22] | PCa (n=36) and benign tissue (n=14). |
| GSE6956 [23] | Primary PCa (n=69) and normal adjacent prostate (n=18) |
| GSE70768 [24] | Primary PCa (n=112), benign tissue (n=74) and CRPC (n=13) |
| TCGA-PRAD [25] | Primary PCa (n=497) and normal adjacent tissue samples (n=51) |
| GSE21034 [26] | Primary PCa (n=131) and metastatic tissue samples (n=19). |

#### Differential Gene Expression Analyses

We used the *limma* package (Linear Models for Microarray Analysis) [27] to study differential gene expression from both microarrays and RNA-sequencing (RNA-seq). In the case of non-normalized data, quantile normalization was applied [27]. For RNA-seq data, the *voom* function in the *limma* package was used for processing [28]. We conducted pair-wise differential expression analyses within each dataset. For each available probe or gene, the fold changes (FC) between conditions were calculated, and expressed as log_2_FC. To correct for multiple testing, we used the Benjamini-Hochberg method to control the type I error, and reported adjusted p-values.

### Survival analyses

#### Dataset selection criteria

To perform survival analysis, we searched Gene Expression Omnibus (GEO), cBioPortal and the Genomic Data Commons Data Portal to identify eligible datasets that met the following criteria: (1) PCa cases with available gene expression data and (2) available clinico-pathological features with ≥5 years of follow-up. Gene expression and clinical data were downloaded and analyzed for the resulting selected datasets. Samples with incomplete gene expression data or missing essential clinico-pathological metadata were not included. Datasets were randomly distributed in training (5 datasets, 7 survival analyses) and validation cohorts (4 datasets) (**Table 2**).

**Table 2:** PCa transcriptomics datasets for survival analyses.

| **<u>Dataset</u>** | **<u>Samples</u>** | **<u>Survival end-point</u>** | **<u>Covariates</u>** | **<u>Cohort</u>** |
| --- | --- | --- | --- | --- |
| TCGA-PRAD [25] | 497 PCa (RNAseq) | Disease Progression<br>Disease-Free Time (n=337) | Gleason Group, PSA levels, Clinical T Stage, Targeted Molecular/ Radiation Therapy | Training |
| GSE70768 [24] | 111 PCa (Microarray) | Biochemical Relapse | Age, Gleason Group, PSA levels, T Stage | Training |
| GSE70769 [24] | 92 PCa (Microarray) | Biochemical Relapse | Gleason Group, PSA levels, T Stage | Training |
| GSE116918 [29] | 248 PCa (Microarray) | Metastasis Development<br>Relapse | Age, Gleason Score, PSA levels, T Stage | Training |
| GSE16560 [30] | 281 PCa (Microarray) | Death | Age, Gleason Group | Training |
| GSE54460 [31] | 106 PCa (RNA-seq) | Biochemical Relapse | Age, Gleason Score, PSA levels, T Stage | Validation |
| GSE94767 [32] | 233 PCa (Microarray) | Biochemical Relapse | Gleason Group, PSA levels, T Stage | Validation |
| DKFZ [33] | 81 PCa (RNA-seq) | Biochemical Relapse | Age, Gleason Score, PSA levels, T Stage | Validation |
| SU2C-PCF [34] | 81 metastatic CRPC (RNA-seq) | Death | Age, Gleason Score, PSA levels | Validation |

#### Survival analyses

We used the log-rank test to analyze differences in the risk of disease progression-events between different groups of patients [35]. To stratify patients according to high-or low-expression, we used the Cutoff Finder tool to find the optimal cutoff point for each gene [36]. The Cox proportional hazards model was used to estimate the risk of the disease progression-event for the different groups [37]. Multivariable analyses included clinico-pathological features as covariables. All modeling, calculations, and graphs were performed with the R packages *survival* [38] and *survminer* [39].

#### Selection of candidate genes for modeling a risk score

To identify the 15 most important genes for predicting events we used a machine learning ensemble based approach (i.e Random Forest Classifier) as implemented in the *randomForestSRC* R package [40]. The *mtry* and *nodesize* parameters were optimized through a grid search approach to minimize the out-of-bag error. We used the Breiman-Cutler variable importance (VIMP) measure to estimate the relative importance of each variable in predicting event-free survival within the training datasets. We applied the subsampling method [41] to estimate the standard error of the VIMP and to calculate the confidence intervals. Genes were ranked according to their variable importance. To facilitate the comparison across datasets, VIMP values were converted into fractions, with 1 representing the most important variable and 0 representing the least important variable within a given dataset.

#### Gene Signature and Risk Score Calculation

We modelled a risk score based on the gene expression of the 15 most important genes identified across training datasets using Random Forest. To develop this risk score, we calculated model coefficients through Lasso regression using TCGA-PRAD data. Patient scores were then calculated based on the expression of the selected genes following Lasso regression. The performance of this risk score was evaluated within each training dataset. Univariable Cox regression was used to estimate the risk of poor survival in patients with high-risk scores. Patients were stratified either by a dichotomized risk score (with the median as the cutpoint) or by a continuous risk score. The concordance index (CI) was used to measure the performance of the signature within each dataset.

In the validation stage, those same coefficients were used in all additional datasets. For each patient, the score was calculated, and its association with event-free survival were studied using univariable and multivariable Cox regressions.

#### Transcriptome analysis of MDA PCa PDXs

To assess the association of the stemness-signature and other clinico-pathological characteristics in an extensively annotated cohort, we used the MDA PCa PDX series, which was previously developed in the “Prostate Cancer Patient Derived Xenograft Program” at MD Anderson Cancer Center and the David H. Koch Center for Applied Research of Genitourinary Cancers [42]. Briefly, PCa tissue samples used were derived from various procedures and small pieces were then implanted into subcutaneous pockets of 6 to 8 week-old male CB17 SCID mice (Charles River Laboratories) [42]. RNA-Seq and transcriptome analysis on these samples was performed as previously described [43].

#### Unsupervised Clustering and Principal Component Analysis (PCA)

Unsupervised clustering analysis including the expression data of the stemness-associated genes included in the signature was performed using the *pheatmap* [44] package and Principal Component Analysis was performed using the *factoextra* package in R [45].

#### Receiver Operating Characteristic (ROC) curve for NEPC classification

*pRoc* package [46] was used for the estimation of receiver operating characteristic (ROC) curve and area under ROC curve (AUC).

#### NEPC patients samples dataset

To assess gene expression in NEPC samples, we downloaded the data from the Neuroendocrine Prostate Cancer (Multi-Institute, Nat Med 2016) dataset published by Beltran *et al.* [12] from cBioPortal [47–49]. Briefly, this dataset contains transcriptomics and histopathological data from 49 PCa samples (34 CRPC-Adeno and 15 CRPC-NE) obtained by RNA-Seq.

#### Statistical analyses

All bioinformatics analyses were performed using the R programming language [50] through the RStudio platform (RStudio, PBC, Boston, MA, USA) [51]. The *tidyverse* package was used for general data analysis and manipulation [52]. For graphics, the packages *ggplot2* [53], *ggpubr* [54], and *RColorBrewer* [55] were used. Datasets available in GEO were downloaded with *GEOquery* [56]. All heatmaps were created with the *pheatmap* package [44]. Forest plots were created using GraphPad Prism (La Jolla, CA, USA). Student’s t test and ANOVA followed by Tukey’ test were used to assess differences in risk score values across groups. We used the log-rank test and Cox proportional hazard model regression to study the association between gene expression and patients’ survival. Multivariable analyses were performed in R and plotted in GraphPad Prism software (La Jolla, CA, USA). Statistical significance was set at p ≤ 0.05.

## RESULTS

### Dysregulation of stemness-associated genes across multiple PCa comparisons

We gathered 144 stemness-associated genes in PCa from literature [6,14–18] (**Supplementary Table 1**) and analyzed their expression and association with multiple survival endpoints (**Figure 1A**). First, we performed pair-wise differential gene expression analyses using 7 PCa datasets (n=1,259), which included 11 comparisons between normal prostate, primary tumor, metastatic and castration-resistant PCa (CRPC) samples (**Figure 1A**). Volcano plots evidenced dysregulation of 139 stemness-associated genes, with both up-(red, adjusted p<0.05, log_2_FC>0) and down-regulation (blue, adjusted p<0.05, log_2_FC<0) in all comparisons (Primary PCa *vs.* Benign/Normal/Adjacent, Metastatic PCa *vs.* Benign, Metastatic PCa *vs.* Primary PCa, CRPC *vs.* Benign; CRPC *vs.* Primary PCa) (**Figure 1Bi**). **Figure 1Bii** summarizes these results across comparisons. We observed that all stemness-genes were dysregulated in at least one dataset, with 29 genes consistently up-regulated and 26 genes consistently down-regulated **(Figure 1Bii, Supplementary Table 2**).

**Figure 1.**
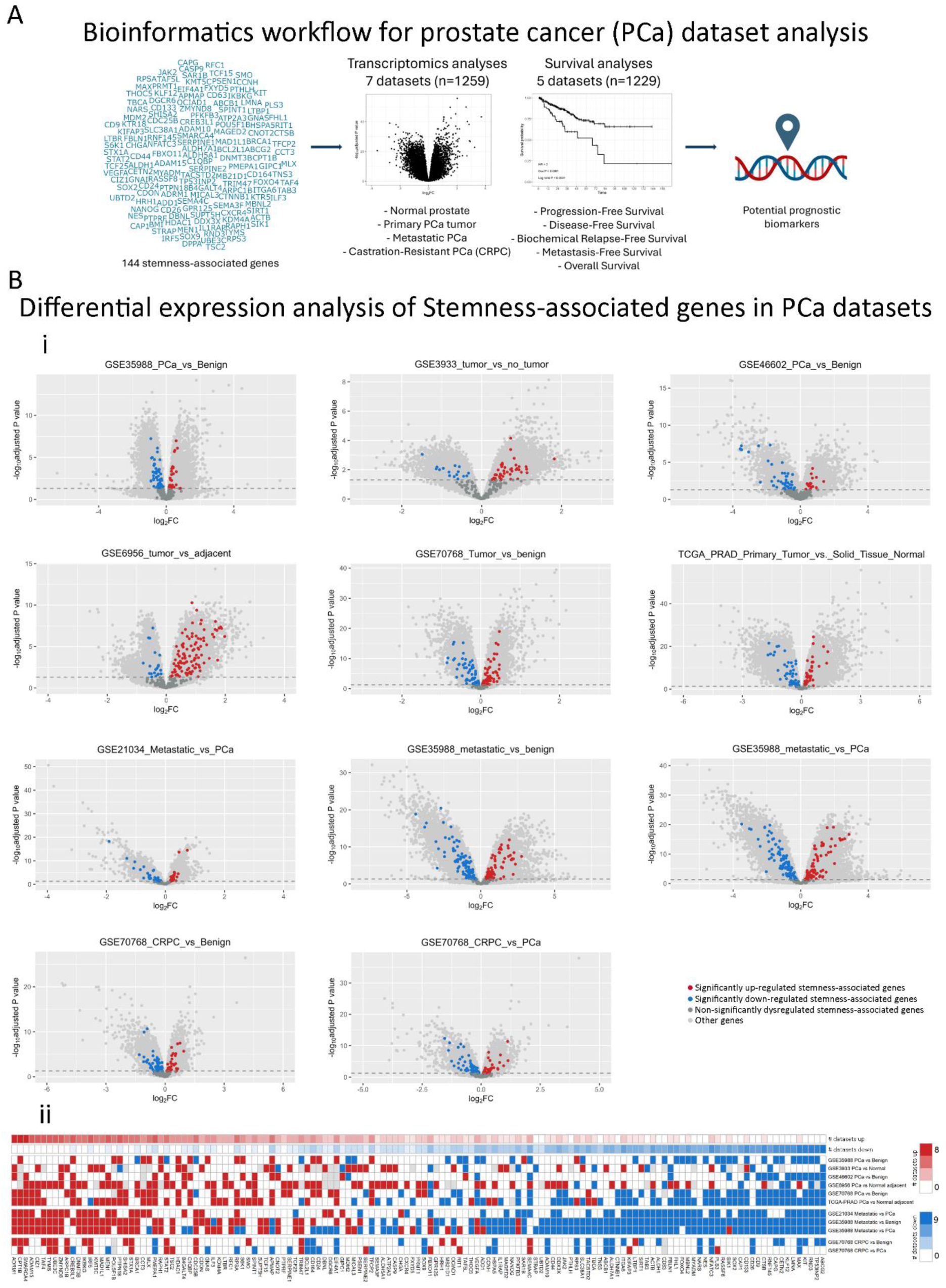
Stemness-associated gene expression changes in PCa patient samples using multiple public datasets. **A)** Schematic representation of gene selection, transcriptomics and survival analyses to define potential prognostic biomarkers. **B) i)** Volcano plots showing the results of the differential expression analysis of all available genes within the included transcriptomics datasets. Red = significantly upregulated stemness-associated gene. Blue = significantly downregulated stemness-associated gene. Dark gray = Non-significantly dysregulated stemness-associated genes. Light gray = other genes available in the dataset. **ii)** Summary heatmap of the transcriptomics analyses performed in multiple publicly available datasets (n=1259). Genes of interest and the results of the differential expression analysis for each dataset are displayed. Each row represents the results of a specific comparison. Annotation depicts the absolute number of comparisons in which each gene is up (red) or downregulated (blue). Red = significantly upregulated gene. Blue = significantly downregulated gene. White = not significant changes. Gray = non available. Datasets: GSE35988 (n=122); GSE3933 (n=103); GSE46602 (n=50); GSE6956 (n=87); GSE70768 (n=179); TCGA-PRAD (n=548); GSE21034 (n=150). Statistical significance was set to adjusted p value<0.05.

### Association of stemness markers with PCa patients’ survival

We evaluated the association with different events in PCa patients, including progression-free survival (PFS), biochemical relapse-free survival (RFS), metastasis-free survival (MFS), overall survival (OS), and disease specific survival (DSS) for the 139 differentially expressed genes. **Figure 2Ai** shows representative Kaplan-Meier plots for three example genes (*DBNL, UBTD2,* and *MBNL2*) in the TCGA-PRAD dataset (n=497, PFS). Results showed that high expression of *DBNL,* and low expression of *MBNL2* were significantly associated with poor PFS (HR=2, Log-rank P=0.0011 and HR=0.39, Log-rank P<0.0001, respectively. **Figure 2Ai**, left and right panels). No significant associations were observed for *UBTD2* (Log-rank P=0.1062, **Figure 2Ai**, middle panel). **Figure 2Aii** shows a heatmap summarizing the results of the univariable survival analysis for each of the 139 candidate genes performed across the 5 training datasets including 5 different types of events (n=1,229; detailed in **Supplementary Table 3**). The results are color-coded as follows: red squares represent genes with high expression significantly associated with shorter times to the event, white squares indicate genes with no significant associations to the event, and blue squares represent genes with high expression significantly associated with a better outcome. Of note, there was a group of genes whose high expression was consistently associated with poor prognosis (**Figure 2Aii**, left, in red), while others were associated with a better outcome (**Figure 2Aii**, right, in blue).

**Figure 2.**
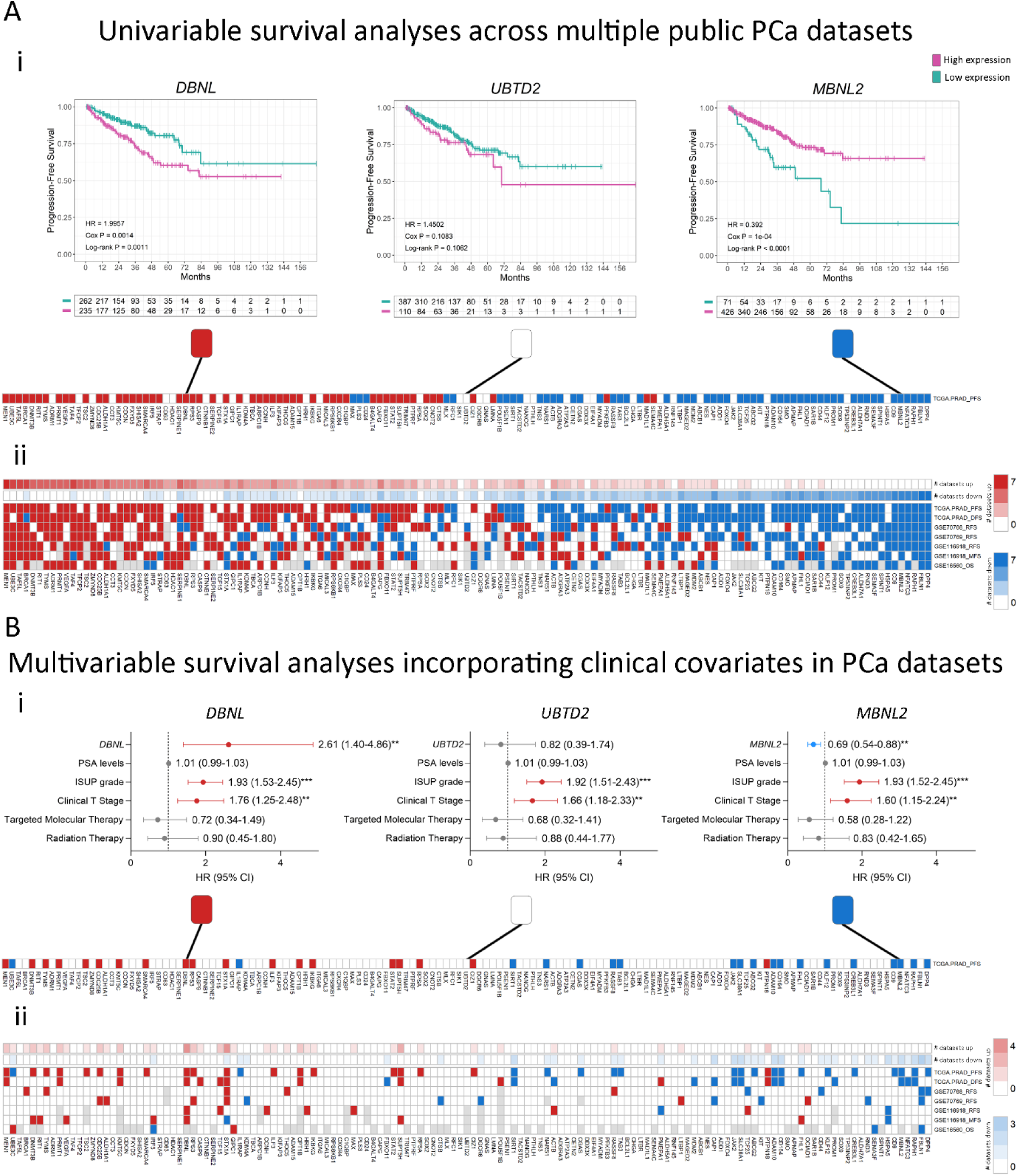
Uni and multivariable survival analysis. A) **i)** Examples of Kaplan-Meier (KM) curves depicting the association of each gene to the risk of event (purple = high expression of a gene; green: low expression of a gene). HR: Hazard Ratio; Cox P: p-value from the Cox proportional hazards model. Log-rank P: p value of the log-rank test. **ii)** Summary heatmap of the univariable survival analyses performed on multiple datasets. The red box indicates that high gene expression is associated with a high risk of an event (HR>1 and Cox P<0.05), blue boxes indicate that high gene expression is associated with a low risk of survival-related events (HR<1 and Cox P<0.05) and white boxes indicate that there are no significant associations between gene expression and risk of an event. Gray = gene no available. Patients were stratified by the median expression of each gene. **B) i)** Examples of forest plots depicting the association of each gene to the risk of event adjusted for all available covariates using the TCGA-PRAD dataset. **ii)** Summary heatmap of the multivariable survival analyses performed on multiple datasets. The red box indicates that high gene expression is associated with a high risk of an event (HR>1 and Cox P<0.05), blue boxes indicate that high gene expression is associated with a low risk of survival-related events (HR<1 and Cox P<0.05) and white boxes indicate that there are no significant associations between gene expression and risk of an event. Gray = gene no available. All comparisons consider low-expression patients as the reference group. Annotation depicts the absolute number of comparisons in which high expression of each gene is associated with high (red) or low (blue) risk. OS: Overall Survival; DSS: Disease-Specific Survival; PFS: Progression-Free Survival; RFS: Relapse-Free Survival; MFS: Metastasis-Free Survival. Datasets: TCGA-PRAD (n=497 PFS, n=337 DFS); GSE70768 (n=111 RFS); GSE70769 (n=92 RFS); GSE116918 (n=248 RFS and MFS); GSE16560 (n=281 OS). Statistical significance was set at Cox P<0.05. **Cox P<0.01; ***Cox P<0.001.

Next, we performed multivariable Cox regression analyses for each of the 139 previously mentioned genes to evaluate their independence from other known risk factors for PCa progression in predicting an event (**Table 2**). For the three examples mentioned above, *DBNL* (HR=2.61, 95% CI 1.40-4.86, Cox P=0.002) and *MBNL2* (HR=0.69, 95% CI 0.54-0.88, Cox P=0.003) displayed a significant association with high and low risk of PFS, respectively, independently from the other covariates available in the TCGA-PRAD dataset (PSA levels, ISUP grade, Clinical T Stage, and Targeted Molecular/Radiation Therapy; **Figure 2Bi**). No significant associations were observed for *UBTD2* (**Figure 2Bi**). The overall results for the multivariable analyses are summarized as a heatmap in **Figure 2Bii** and detailed in **Supplementary Table 4**. Most associations observed in the univariable analysis (**Figure 2Aii**) lost statistical significance after adjusting for clinical covariates (**Figure 2Bii**).

### Modeling a stemness-associated signature with prognostic value

We used a machine learning algorithm to identify the most relevant prognostic candidate genes to model a gene-expression signature that could stratify patients into risk groups of disease progression and death. We used a Random Forest algorithm to rank genes according to their relevance for event prediction in the training datasets and calculated the mean relative importance score for each gene (**Figure 3A**). The top 15 genes were: *ALDH1A1*, *KMT5C*, *DPP4*, *RPS6KB1*, *TYMS*, *CCT3*, *IL1RAP*, *MICAL3*, *CDC25B*, *IRF5*, *MEN1*, *DNMT3B*, *CD24*, *RND3*, and *CASP9* (**Figure 3A**, purple square). Next, we used these genes to develop our stemness-associated risk signature. Model coefficients were calculated on the TCGA-PRAD cohort by Lasso regression, a feature selection method that keeps the most important predictors by shrinking the coefficients of less significant genes to zero. This analysis resulted in a signature of 7 significant genes, generating the following weighted linear model: 0.284×*KMT5C* + 0.2723×*MEN1* + 0.2178×*TYMS* + 0.09×*IRF5* + 0.0827×*DNMT3B* + 0.048×*CDC25B* – 0.0597×*DPP4*, where gene expressions are considered a continuous variable (**Figure 3Bi**).

**Figure 3.**
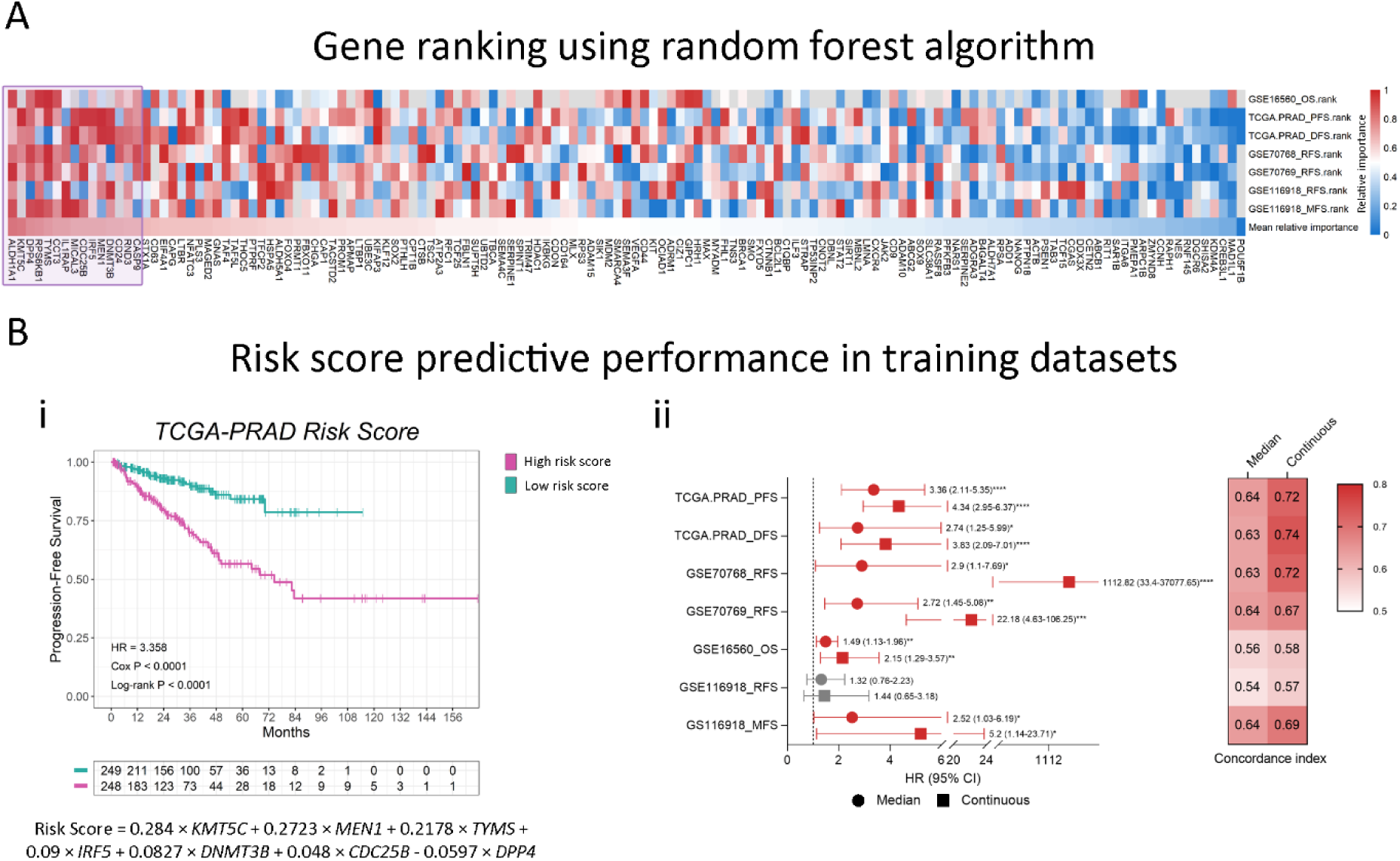
Machine learning Random Forest algorithm for prognostic candidates’ selection. **A)** Heatmap summarizing the relative importance of the variables (genes) for all training datasets. The relative importance was converted into percentiles, where 1 represents maximum relative importance (red) and 0 indicates minimum relative importance (blue). Gray = gene not available in the dataset. The 15 top-ranked genes (purple) were selected as candidates for our stemness-associated risk signature. **B) i)** Example of Kaplan-Meier (KM) curve using the TCGA-PRAD dataset depicting the association of the 7-gene score to the risk of progression (purple = high 7-gene score; green: low 7-gene score). The coefficients for each gene were calculated by Lasso regression using TCGA-PRAD data, and the 7-gene score was constructed as follows: 0.284×*KMT5C* – 0.0597×*DPP4* + 0.2178×*TYMS* + 0.048×*CDC25B* + 0.09×*IRF5* + 0.2723×*MEN1* + 0.0827×*DNMT3B*. Patients were stratified by the median of the score. HR: Hazard Ratio; p-value: p-value from the Cox proportional hazards model. Log-rank P: p value of the log-rank test. **ii)** Summary forest plot displaying the survival analysis of the association of the 7-gene signature with the risk of disease progression-events in the training datasets. Patients survival was analysed by either stratification by the median of the 7-gene score (circles) or taking the 7-gene score as a continuous variable (squares). On the right, heatmap depicting the concordance index value for each of the analyses. The concordance index is a performance measure of the signature within each dataset. Cox P = p-value of the Cox regression coefficient. HR = Hazard Ratio. (95% CI) = 95% Confidence Interval. PFS: Progression-Free Survival; DFS: Disease-Free Survival; RFS: Relapse-Free Survival; OS: Overall Survival; MFS: Metastasis-Free Survival. Statistical significance was set at Cox P<0.05. *Cox P<0.05; **Cox P<0.01; ***Cox P<0.001; ****Cox P<0.0001.

Next, in order to evidence this 7-gene signature prognostic performance, we calculated the risk score for each patient on the TCGA-PRAD dataset and stratified them into high-risk and low-risk groups using the median score as the cutpoint. As expected, we evidenced a shorter progression time for the high-risk group compared to the low-risk group (HR=3.36, 95% CI 2.11-5.35, Log-rank P<0.0001, **Figure 3Bi**). When we considered the score as a continuous variable, we observed a HR=4.34 (95% CI 2.95-6.37, Cox P<0.0001) for each unit increase in the score (**Figure 3Bii**, **Supplementary Table S5**). We then extended the score to the other training datasets and corroborated its prognostic significance within these cohorts. When analyzing the event-free survival in all the other training datasets, we observed significant associations with our model in 5 out of 6 analyses, both using the dichotomous and continuous score, suggesting our 7-gene signature is able to predict risk of multiple disease progression-events across our training cohorts (**Figure 3Bii**, **Supplementary Table S5**). The identified genes and the developed risk score model effectively stratify patients based on their risk of adverse outcomes, suggesting their potential as prognostic biomarkers.

### Consistent performance across validation datasets

Next, we validated our model using datasets from independent cohorts (n=501). We calculated the 7-gene score for all patients in the different datasets and categorized them into high or low risk using the median as a cutoff. Interestingly, the risk score was significantly associated with event-free survival in all validation cohorts (**Figure 4Ai-ii**). Of note, in the SU2C dataset, which comprises metastatic PCa samples, patients with high score had near 2-fold risk of death compared to patients with low score (**Figure 4Aii**). This demonstrates that the 7-gene signature is a robust predictor of the risk of death even in advanced stages. Moreover, when analyzing the 7-gene signature as a continuous variable, all datasets presented significant results, with higher concordance indexes than the dichotomized analysis (**Figure 4Aiii**). Multivariable analyses demonstrated that our score predicts disease progression-events independently of the other clinico-pathological variables (**Figure 4B**), which highlights its potential utility in clinical decision-making.

**Figure 4.**
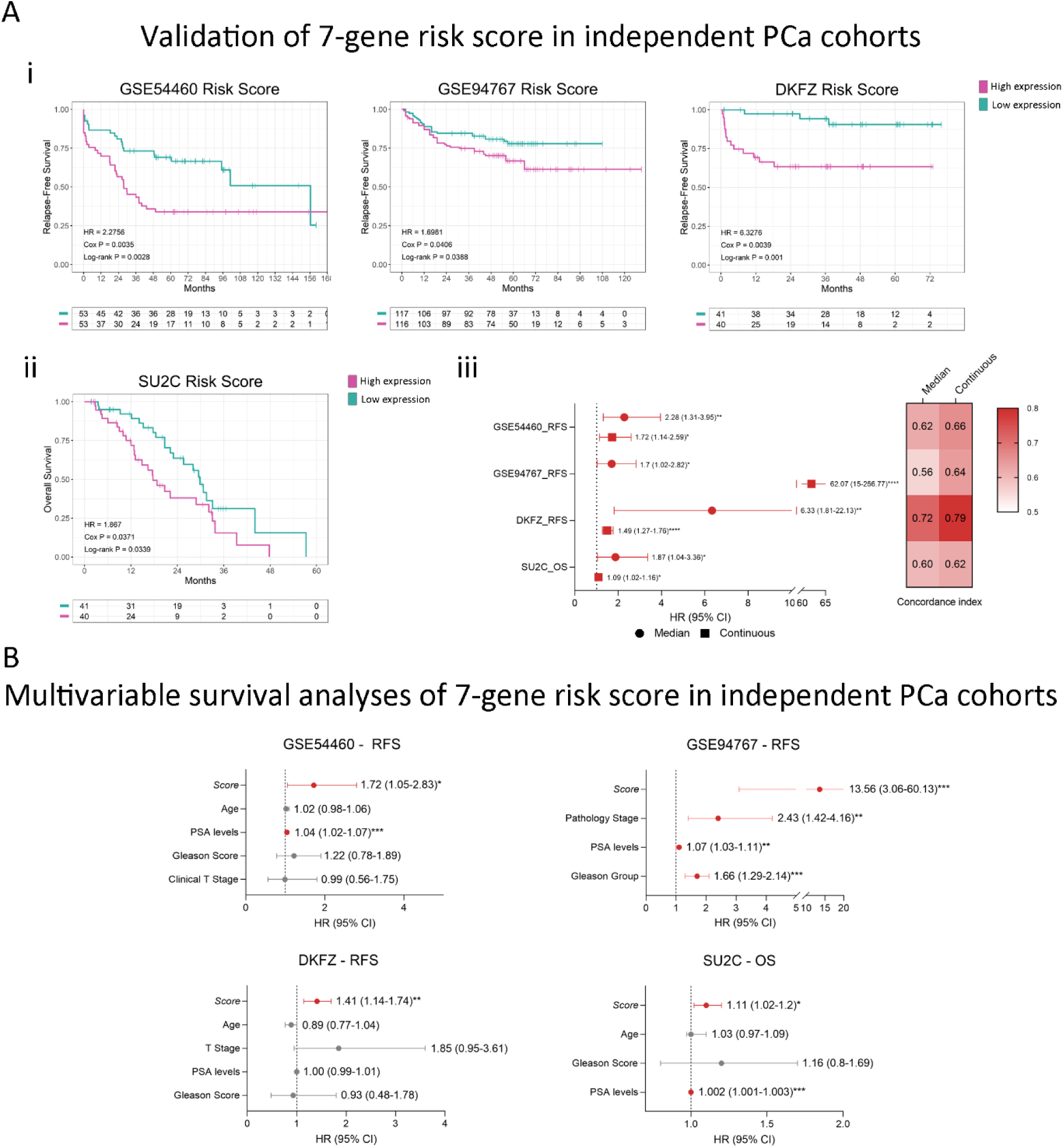
Gene signature’s performance across external validation datasets A) **i)** Kaplan-Meier curves depicting the association of the 7-gene score to the risk of disease progression-events included in the validation datasets. The coefficients for each gene were calculated by Lasso regression using TCGA-PRAD data, and the 7-gene score was calculated as follows: 0.284×*KMT5C* + 0.2723×*MEN1* + 0.2178×*TYMS* + 0.09×*IRF5* + 0.0827×*DNMT3B* + 0.048×*CDC25B* – 0.0597×*DPP4*. Patients were stratified by the median of the score. HR: Hazard Ratio; Cox P: p-value from the Cox proportional hazards model. Log-rank P: p value of the log-rank test. **ii)** Summary forest plot displaying the survival analysis of the association of the 7-gene signature with the risk of disease progression-events in the validation datasets. Patients survival was analysed by either stratification by the median of the 7-gene score (circles) or taking the 7-gene score as a continuous variable (squares). On the right, heatmap depicting the concordance index value for each of the analyses. The concordance index (CI) is a performance measure of the signature within each dataset. RFS: Relapse-Free Survival; OS: Overall Survival. **B)** Forest plots depicting the association of each gene to the risk of event adjusted for all available covariates within each validation dataset. Cox P = p-value of the Cox regression coefficient. HR = Hazard Ratio. [95% CI] = 95% Confidence Interval. Datasets: GSE54460 (n=106); GSE94767 (n=233); DKFZ (n=81); SU2C-PCF (n=81).Statistical significance was set at Cox P<0.05. *Cox P<0.05; **Cox P<0.01; ***Cox P<0.001; ****Cox P<0.0001.

### The stemness-associated gene signature captures neuroendocrine disease heterogeneity in the MDA PCa PDX series

Next, we sought to analyze the association between the 7-gene signature and other clinico-pathological characteristics available in the MDA PCa PDX series, which was developed in the Laboratory of Dr. Navone within the “Prostate Cancer Patient Derived Xenograft Program” at MD Anderson Cancer Center and the David H. Koch Center for Applied Research of Genitourinary Cancers. PCa tissue samples used for PDX development were derived from therapeutic or diagnostic procedures, namely, radical prostatectomies, orthopedic, and neurosurgical procedures to palliate complications, and biopsies of metastatic lesions [42] (**Figure 5A**). We analyzed the expression of the 7 stemness-associated genes selected in the present study using previously generated RNA-Seq data from the 44 MDA PCa PDXs [43]. Surprisingly, the expression of this signature was able to accurately cluster PDXs according to their histopathological classification (adenocarcinoma or sarcomatoid vs. neuroendocrine tumors) in an unsupervised clustering analysis (**Figure 5Bi**). Moreover, NEPC PDXs displayed significantly higher scores (**Figure 5Bii**). Specifically, *CDC25B*, *TYMS*, *KMT5C* and DNMT3B were significantly upregulated in NEPC vs. no-NEPC PDXs, while *IRF5* and *DPP4* were significantly downregulated (**Figure 5Biii**).

**Figure 5.**
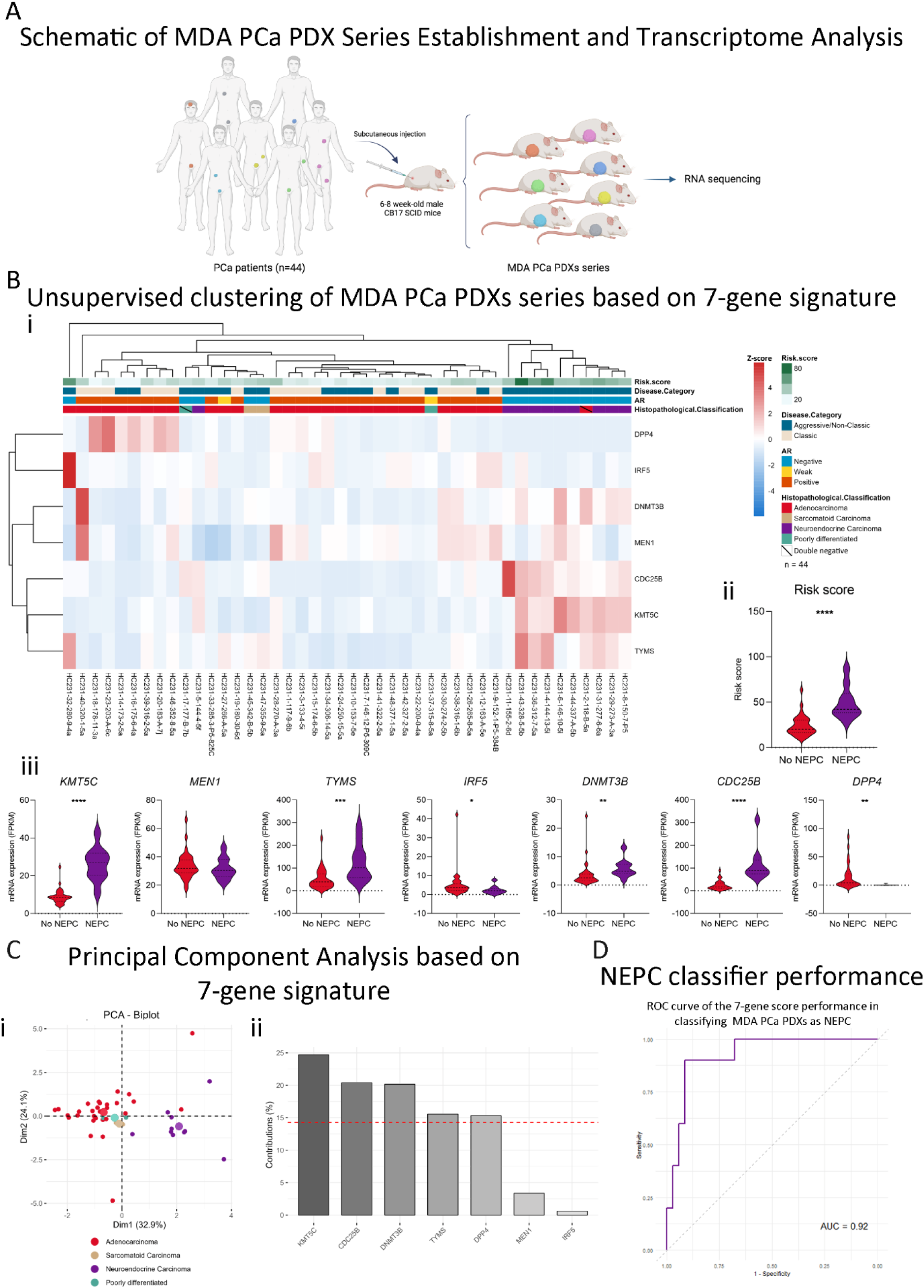
Transcriptome analysis of the MDA PCa PDX series. **A)** Schematic representation of the MDA PCa PDX series establishment and transcriptome analysis (n=44) (created with BioRender.com). **B) i)** Heatmap depicting unsupervised clustering analysis of RNAseq data from the 44 MDA PCa PDXs considering the expression of the 7-gene signature (*KMT5C, MEN1, TYMS, IRF5, DNMT3B, CDC25B and DPP4*). Red, white, and blue represent greater, intermediate, and lower gene expression levels. **ii)** Violin plot showing the 7-gene score levels in no-NEPC and NEPC samples from the MDA PCa PDX series. **iii)** Violin plots showing the expression levels (FPKM) of the genes included in the 7-gene score in no-NEPC and NEPC samples from the MDA PCa PDX series. **C) i)** PCA biplot considering the expression of the 7-gene signature using the MDA PCa PDX data assessed by RNA-seq. Each point represents one PDX. Samples are coloured according to the histopathological classification: adenocarcinoma (red), sarcomatoid (beige) and neuroendocrine (purple). **ii)** Bar plot showing the contribution (%) of each gene in the signature to the variance in the PC1 from the PCA. **D)** ROC curve showing the performance of the 7-gene score in classifying MDA PCa PDXs as NEPC. Statistical significance was calculated using Student’s t test and was set at p<0.0.5. *p<0.05; **p<0.01; ***p<0.001; ****p<0.0001.

These results were also observed in a Principal Component Analysis (**Figure 5Ci**), which highlighted *KMT5C* as the main gene in the signature contributing to the variance (PC1) between samples of different histopathological profiles (**Figure 5Cii**), followed by *CDC25B* and *DNMT3B*. Of note, *KMT5C* is also the gene that weighs higher in our score (**Figure 3Ci**). To evaluate the power of the signature in predicting whether a tumor is NEPC, we performed ROC analysis. The AUC of our 7-gene score was 0.92 (**Figure 5D**), highlighting its high performance for classifying NEPC samples.

### Our stemness-score adds value to pre-existing NEPC score

To compare our risk score performance with a pre-established NEPC classification score, we analyzed the expression of the genes from the 70-gene signature by Beltran et al. [12], in the MDA PCa PDX series. We observed a good segregation of the PDXs according to their histopathological classification when using the 70 genes from Beltran *et al.* NEPC score; however, the 2 double-negative tumors (negative for AR and NE features) were clustered within the NEPC tumors group (**Figure 6Ai**). Nonetheless, when also including the expression of the 7 genes identified in this work alongside the genes from the NEPC classification score [12], clustering of the PDX was more accurate, not only grouping adenocarcinomas vs. NEPC tumors, but also sarcomatoid samples (**Figure 6Aii**).

**Figure 6.**
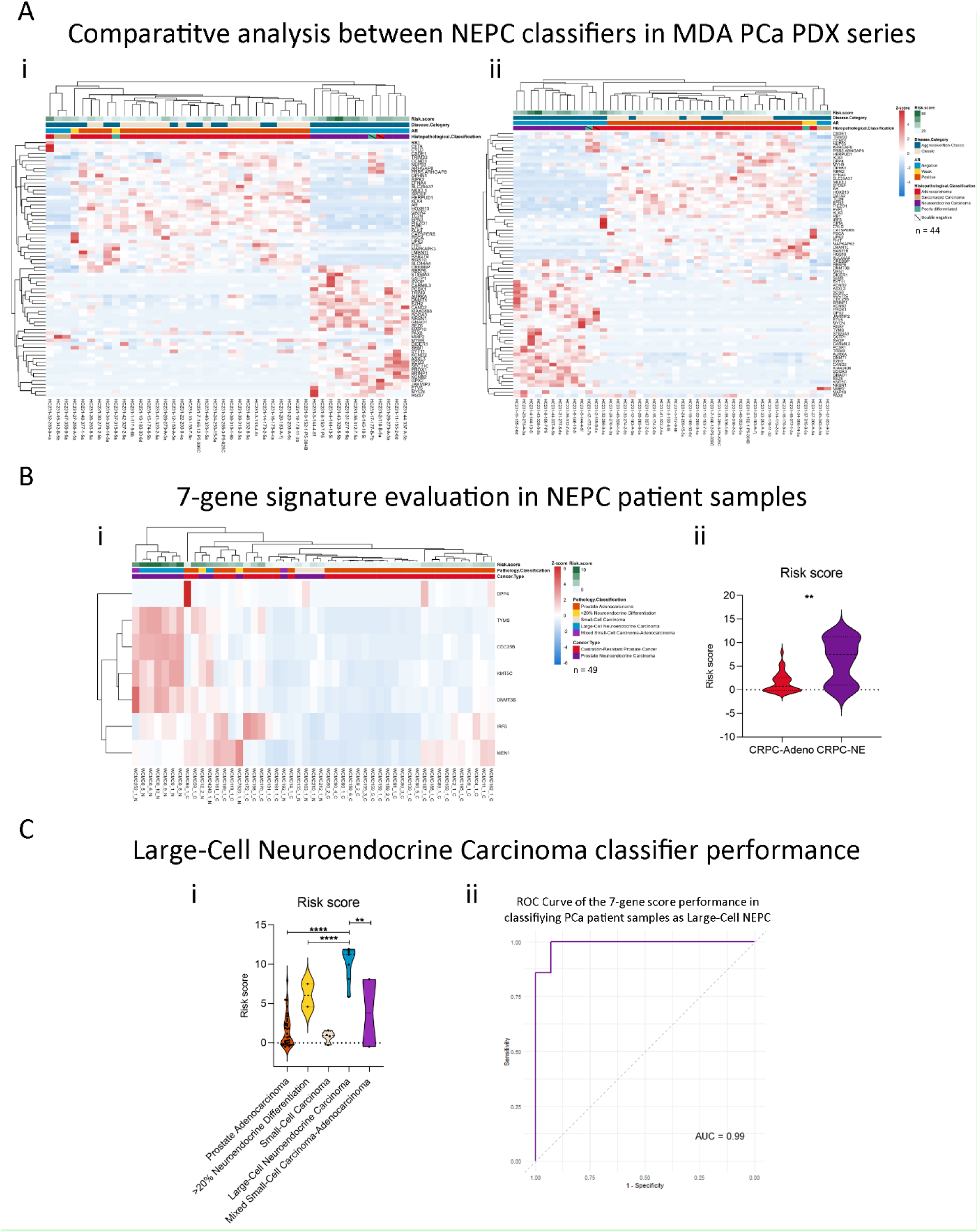
Clinical validation in NEPC samples. A) **i)** Heatmap depicting an unsupervised clustering analysis of RNAseq data from the MDA PCa PDX series considering the expression of the 70-gene signature proposed by Beltran *et al.* [12] **ii)** Heatmap depicting an unsupervised clustering analysis of RNAseq data from the MDA PCa PDX series considering the expression of the 70-gene signature proposed by Beltran *et al*. plus the 7 genes (*KMT5C, MEN1, TYMS, IRF5, DNMT3B, CDC25B and DPP4*) from the risk score model propose in our work. **B) i)** Heatmap depicting an unsupervised clustering analysis of RNAseq data from human patients in Beltran *et al.*, dataset (n=49) [12] considering the expression of the 7-gene signature. Red, white, and blue represent greater, intermediate, and lower gene expression levels. Expression values are presented as z-scores. **ii)** Violin plot showing 7-gene score levels in CRPC-Adeno and CRPC-NE samples from the Beltran *et al.*, dataset. **C) i)** Violin plot showing risk score levels in samples from the Beltran *et al.*, dataset according to the histological classification: prostate adenocarcinoma without neuroendocrine differentiation, prostate adenocarcinoma with neuroendocrine differentiation >20%, small-cell carcinoma, large-cell neuroendocrine carcinoma, and mixed small-cell carcinoma–adenocarcinoma. **ii)** ROC curve showing the performance of the 7-gene score in classifying PCa patient samples from Beltran *et al.* dataset as Large-Cell NEPC. Statistical significance was calculated using Student’s t test or ANOVA followed by Tukey’s test, and was set at p<0.05. **p<0.01; ****p<0.0001.

### The 7-gene signature effectively classifies large-cell neuroendocrine carcinomas

To validate the association of our risk score model with NEPC, we analyzed the transcriptomics dataset from Beltran *et al.* (n=49) [12], which includes 15 samples from CRPC-neuroendocrine (NE) and 34 CRPC-adenocarcinomas tumors. Our signature was able to distinguish CRPC-NE tumors to a limited extent (**Figure 6Bi**), while, overall, our risk score was significantly higher in CRPC-NE compared to CRPC-Adeno (p<0.01, **Figure 6Bii**). However, we looked further into the available pathology classification (prostate adenocarcinoma with no neuroendocrine differentiation, n=34; prostate adenocarcinoma with neuroendocrine differentiation >20%, n=2; small-cell carcinoma n=4; large-cell neuroendocrine carcinoma, n=7; mixed small-cell carcinoma– adenocarcinoma, n=2) and observed that 6/7 samples of the large-cell NEPC clustered together (**Figure 6Bi**), while the 7-gene signature was particularly higher in that subtype (**Figure 6Ci**). Strikingly, the AUC=0.99 suggests that the signature of 7 stemness-associated genes proposed in this work is accurate in classifying samples as large-cell NEPC (**Figure 6Cii**). Since large-cell NEPC molecular characterization remains elusive [57], our findings set ground for future research on the implications of these genes in this subtype pathogenesis.

## DISCUSSION

In this study we identified and validated a novel 7-gene signature that represents a significant advancement in the prediction of poor outcomes and molecular detection of NEPC. Our findings demonstrate that this signature not only reliably stratifies PCa patients based on their risk of progression but also reveals a crucial link between stemness-associated pathways and neuroendocrine characteristics. Importantly, this signature is particularly adept at identifying tumors within the Prostate Cancer Foundation (PCF) and World Health Organization (WHO)-defined large-cell neuroendocrine carcinoma [58,59], which is regarded as very rare and associated with very poor outcomes (mean survival of 7 months) [59].

Large-cell NEPC are high grade tumors that usually develop from treatment-resistant clones [60]; they are mainly diagnosed histopathologically, thus remaining a challenge and underrecognized [57,58,61]. Hence, there is a need for molecular biomarkers that could subclassify NEPC tumors for better clinical management [57]. The ability of our 7-gene signature to pinpoint this specific aggressive and challenging NEPC subtype underscores the clinical utility of our model in guiding more precise therapeutic interventions.

Our stemness-associated signature addresses a critical need for improving PCa prognosis, while also offering precise stratification of NEPC, which is often characterized by poor clinical outcomes and high proliferative indices [62]. NEPC is recognized as one of the most aggressive and treatment-resistant forms of PCa, often arising in the context of advanced CRPC after multiple rounds of ADT [63]. While most NEPC cases develop in patients with a history of extensive anti-androgen treatment, the disease can also manifest *de novo*, albeit rarely, in treatment-naïve patients [9,12]. Further, ADT-induced NE transdifferentiation could be explained by altered mast cell infiltration [64,65]. Maimaitiyiming *et al.* have established a mast cell gene signature with prognostic efficacy in PCa [66], and, interestingly, mast cells have been reported to support the stem phenotype of cancer cells [67]. Altogether, focusing on stemness-associated genes could offer insights into NEPC biology and potential targets.

The molecular landscape of NEPC has been increasingly clarified in recent years, with significant contributions from studies like those of Beltran *et al.*, who have delineated the heterogeneity within NEPC and highlighted distinct molecular subtypes [12,58,68]. Their research highlights the genetic, epigenetic and molecular diversity of NEPC, particularly noting alterations such as RB1 and TP53 loss, MYCN overexpression, and the activation of the PI3K/AKT pathway, which contribute to the aggressive nature of these tumors [12,58,68]. Our study builds on these findings by focusing on a 7-gene stemness signature. Unlike previous signatures that include a broad array of genes, our streamlined 7-gene model achieves comparable or superior predictive accuracy, underscoring its practical utility in diverse clinical contexts.

The biological relevance of the genes in our signature — *KMT5C*, *MEN1*, *TYMS*, *IRF5*, *DNMT3B*, *CDC25B* and *DPP4* (also known as *CD26*)— lies in their involvement in critical processes such as chromatin modification, DNA methylation, DNA repair, cell cycle regulation, immune escape and extracellular matrix remodelling [69–75]. These processes are fundamental to maintaining the plasticity and adaptability of cancer stem cells (CSCs) [5,76], which are enriched after transdifferentiation of prostate adenocarcinoma into more aggressive neuroendocrine phenotypes [77]. For example, *KMT5C, DNMT3B* and *MEN1* play pivotal roles in chromatin remodeling and methylation [69–71], processes that are crucial for the epigenetic reprogramming observed in NEPC [78]. Additionally, *TYMS* has been previously associated with neuroendocrine differentiation in other types of cancer [79,80]. The integration of these stemness-associated genes into our model highlights the potential for characterizing NEPC-like tumors.

One of the key strengths of our study is the extensive validation of our signature across patient datasets and PDXs models. The latter, which faithfully replicate the histological and genetic features of human tumors, are widely regarded as the gold standard for preclinical studies [81]. Our findings demonstrate that the 7-gene signature consistently distinguishes NEPC from other PCa subtypes in these models, underscoring its clinical utility and its potential for identifying NEPC-like tumors. This aspect of our research not only validates the predictive power of the signature but also highlights its potential utility in translational research, particularly in the development of novel therapeutic strategies aimed at targeting the molecular underpinnings of NEPC.

## LIMITATIONS

Despite the robustness of our findings, there are several limitations that must be acknowledged. Our study primarily relies on transcriptomic data from publicly available repositories, while comprehensive, may not fully represent the genetic diversity of PCa patients globally. Future research should focus on further validating our signature in ethnically and genetically diverse cohorts to ensure its broad applicability. Additionally, while our focus on transcriptomic data has provided valuable insights into NEPC biology, integrating multi-omics data, including proteomics and metabolomics, could enhance the predictive power of our model. Moreover, the scarce number of NEPC samples with transcriptomics data and, particularly, of large-cell NEPC (probably due to under-recognition and underreporting [57]) requires further validation in larger cohorts. Functional validation of the identified genes through *in vivo* studies will also be critical for determining their role in disease and translating findings into clinical practice.

## CONCLUSION

This study presents a significant advancement in PCa prognosis and classification of NEPC, particularly for the challenging large-cell subtype. Importantly, PCa cases presenting this molecular signature, even when not histopathologically identified as NEPC, also exhibit a poor prognosis. This reinforces the clinical relevance of our model, which is capable of identifying aggressive tumor subtypes that may not yet display overt NE differentiation but still represent a high risk for adverse outcomes. Through the development of this novel stemness-associated 7-gene signature, our model offers a robust and practical tool with potential clinical application, paving the way for more personalized and effective therapeutic strategies in PCa.

## Data Availability

All data produced in the present study are available upon reasonable request to the authors.

## ACKNOWLEDGEMENTS

<u>Funding:</u> The present study was supported by Agencia Nacional de Promoción de la Investigación el Desarrollo Tecnológico y la Innovación (ANPCyT) PICT-RAICES-2021-III-A-00080; David H. Koch Center for Applied Research in Genitourinary Cancers at MD Anderson (Houston, TX); and NIH/NCI U01 CA224044. The funders of the study had no role in study design, data collection, data analysis, data interpretation, writing or decision to submit.

## AUTHOR CONTRIBUTIONS

Conceptualization: AS, PS, EV, DA, FC, MM, JC, AT, EL, JB and GG.

Methodology: AS, PS, RS, GP, MM, JC, JB and GG.

Software: AS, PS, MM, JC, and JB.

Validation: AS, PS, RS, GP, EV, MM, JC, AT, JB and GG.

Formal Analysis: AS, PS, NA, DA, FC, EV, MM, JC, AT, JB and GG.

Investigation: AS, PS, RS, GP, AT, JB and GG.

Resources: EV, JC, AT, EL and GG. Data Curation: AS, PS and JB.

Writing – Original Draft Preparation: AS, PS, NA, DA, FC, EV, MM, JC, AT, EL, JB and GG.

Writing – Review & Editing: AS, PS, NA, DA, FC, EV, MM, JC, AT, EL, JB and GG.

Visualization: AS, JB, GG.

Supervision: EV, MM, JC, AT, JB and GG.

Project Administration: JB and GG.

Funding Acquisition: EV, JC, AT, EL and GG.

## SUPPLEMENTARY TABLES

**Supplementary Table S1.** List of 144 stemness-associated genes gathered from PCa literature.

**Supplementary Table S2.** Results of differential expression analyses of the 144 stemness-associated genes across transcriptomics datasets.

**Supplementary Table S3.** Results of univariable survival analyses of the 139 dysregulated stemness-genes.

**Supplementary Table S4.** Results of multivariable survival analyses of the 139 dysregulated stemness-genes.

**Supplementary Table S5.** Score levels across training and validation datasets.

## REFERENCES

1. Global Cancer Statistics 2022: GLOBOCAN Estimates of Incidence and Mortality Worldwide for 36 Cancers in 185 Countries – Bray – 2024 – CA: A Cancer Journal for Clinicians – Wiley Online Library Available online: https://acsjournals.onlinelibrary.wiley.com/doi/10.3322/caac.21834 (accessed on 18 July 2024).

2. Beltran, H.; Rickman, D.S.; Park, K.; Chae, S.S.; Sboner, A.; MacDonald, T.Y.; Wang, Y.; Sheikh, K.L.; Terry, S.; Tagawa, S.T.;, et al. Molecular Characterization of Neuroendocrine Prostate Cancer and Identification of New Drug Targets. Cancer Discov. 2011, 1, 487–495, doi:10.1158/2159-8290.CD-11-0130.

3. Robinson, D.; Van Allen, E.M.; Wu, Y.-M.; Schultz, N.; Lonigro, R.J.; Mosquera, J.-M.; Montgomery, B.; Taplin, M.-E.; Pritchard, C.C.; Attard, G.;, et al. Integrative Clinical Genomics of Advanced Prostate Cancer. Cell 2015, 161, 1215–1228, doi:10.1016/j.cell.2015.05.001.

4. Liu, C.; Kelnar, K.; Liu, B.; Chen, X.; Calhoun-Davis, T.; Li, H.; Patrawala, L.; Yan, H.; Jeter, C.; Honorio, S.;, et al. The microRNA miR-34a Inhibits Prostate Cancer Stem Cells and Metastasis by Directly Repressing CD44. Nat. Med. 2011, 17, 211–215, doi:10.1038/nm.2284.

5. Al Salhi, Y.; Sequi, M.B.; Valenzi, F.M.; Fuschi, A.; Martoccia, A.; Suraci, P.P.; Carbone, A.; Tema, G.; Lombardo, R.; Cicione, A.;, et al. Cancer Stem Cells and Prostate Cancer: A Narrative Review. Int. J. Mol. Sci. 2023, 24, 7746, doi:10.3390/ijms24097746.

6. Maitland, N.J.; Collins, A.T. Prostate Cancer Stem Cells: A New Target for Therapy. J. Clin. Oncol. 2008, 26, 2862–2870, doi:10.1200/JCO.2007.15.1472.

7. Banerjee, P.; Kapse, P.; Siddique, S.; Kundu, M.; Choudhari, J.; Mohanty, V.; Malhotra, D.; Gosavi, S.W.; Gacche, R.N.; Kundu, G.C. Therapeutic Implications of Cancer Stem Cells in Prostate Cancer. Cancer Biol. Med. 2023, 20, 401–420, doi:10.20892/j.issn.2095-3941.2022.0714.

8. Beltran, H.; Tomlins, S.; Aparicio, A.; Arora, V.; Rickman, D.; Ayala, G.; Huang, J.; True, L.; Gleave, M.E.; Soule, H.;, et al. Aggressive Variants of Castration-Resistant Prostate Cancer. Clin. Cancer Res. Off. J. Am. Assoc. Cancer Res. 2014, 20, 2846–2850, doi:10.1158/1078-0432.CCR-13-3309.

9. Aggarwal, R.; Huang, J.; Alumkal, J.J.; Zhang, L.; Feng, F.Y.; Thomas, G.V.; Weinstein, A.S.; Friedl, V.; Zhang, C.; Witte, O.N.;, et al. Clinical and Genomic Characterization of Treatment-Emergent Small-Cell Neuroendocrine Prostate Cancer: A Multi-Institutional Prospective Study. J. Clin. Oncol. 2018, 36, 2492–2503, doi:10.1200/JCO.2017.77.6880.

10. Dardenne, E.; Beltran, H.; Benelli, M.; Gayvert, K.; Berger, A.; Puca, L.; Cyrta, J.; Sboner, A.; Noorzad, Z.; MacDonald, T.;, et al. N-Myc Induces an EZH2-Mediated Transcriptional Program Driving Neuroendocrine Prostate Cancer. Cancer Cell 2016, 30, 563–577, doi:10.1016/j.ccell.2016.09.005.

11. Mu, P.; Zhang, Z.; Benelli, M.; Karthaus, W.R.; Hoover, E.; Chen, C.-C.; Wongvipat, J.; Ku, S.-Y.; Gao, D.; Cao, Z.;, et al. SOX2 Promotes Lineage Plasticity and Antiandrogen Resistance in TP53– and RB1-Deficient Prostate Cancer. Science 2017, 355, 84–88, doi:10.1126/science.aah4307.

12. Beltran, H.; Prandi, D.; Mosquera, J.M.; Benelli, M.; Puca, L.; Cyrta, J.; Marotz, C.; Giannopoulou, E.; Chakravarthi, B.V.S.K.; Varambally, S.;, et al. Divergent Clonal Evolution of Castration-Resistant Neuroendocrine Prostate Cancer. Nat. Med. 2016, 22, 298–305, doi:10.1038/nm.4045.

13. Bluemn, E.G.; Coleman, I.M.; Lucas, J.M.; Coleman, R.T.; Hernandez-Lopez, S.; Tharakan, R.; Bianchi-Frias, D.; Dumpit, R.F.; Kaipainen, A.; Corella, A.N.;, et al. Androgen Receptor Pathway-Independent Prostate Cancer Is Sustained through FGF Signaling. Cancer Cell 2017, 32, 474–489.e6, doi:10.1016/j.ccell.2017.09.003.

14. Huang, R.; Wang, S.; Wang, N.; Zheng, Y.; Zhou, J.; Yang, B.; Wang, X.; Zhang, J.; Guo, L.; Wang, S.;, et al. CCL5 Derived from Tumor-Associated Macrophages Promotes Prostate Cancer Stem Cells and Metastasis via Activating β-Catenin/STAT3 Signaling. Cell Death Dis. 2020 114 2020, 11, 1–20, doi:10.1038/s41419-020-2435-y.

15. Sharpe, B.; Beresford, M.; Bowen, R.; Mitchard, J.; Chalmers, A.D. Searching for Prostate Cancer Stem Cells: Markers and Methods. Stem Cell Rev. Rep. 2013, 9, 721– 730, doi:10.1007/s12015-013-9453-4.

16. Maitland, N.J.; Frame, F.M.; Polson, E.S.; Lewis, J.L.; Collins, A.T. Prostate Cancer Stem Cells: Do They Have a Basal or Luminal Phenotype? Horm. Cancer 2011, 2, 47– 61, doi:10.1007/s12672-010-0058-y.

17. Leong, K.G.; Wang, B.E.; Johnson, L.; Gao, W.Q. Generation of a Prostate from a Single Adult Stem Cell. Nature 2008, 456, 804–810, doi:10.1038/nature07427.

18. Goldstein, A.S.; Huang, J.; Guo, C.; Garraway, I.P.; Witte, O.N. Identification of a Cell of Origin for Human Prostate Cancer. Science 2010, 329, 568–571, doi:10.1126/science.1189992.

19. Grasso, C.S.; Wu, Y.M.; Robinson, D.R.; Cao, X.; Dhanasekaran, S.M.; Khan, A.P.; Quist, M.J.; Jing, X.; Lonigro, R.J.; Brenner, J.C.;, et al. The Mutational Landscape of Lethal Castration-Resistant Prostate Cancer. Nature 2012, 487, 239–243, doi:10.1038/nature11125.

20. Lapointe, J.; Li, C.; Higgins, J.P.; Van De Rijn, M.; Bair, E.; Montgomery, K.; Ferrari, M.; Egevad, L.; Rayford, W.; Bergerheim, U.;, et al. Gene Expression Profiling Identifies Clinically Relevant Subtypes of Prostate Cancer. Proc. Natl. Acad. Sci. U. S. A. 2004, 101, 811–816, doi:10.1073/pnas.0304146101.

21. Malhotra, S.; Lapointe, J.; Salari, K.; Higgins, J.P.; Ferrari, M.; Montgomery, K.; van de Rijn, M.; Brooks, J.D.; Pollack, J.R. A Tri-Marker Proliferation Index Predicts Biochemical Recurrence after Surgery for Prostate Cancer. PLoS ONE 2011, 6, doi:10.1371/journal.pone.0020293.

22. Mortensen, M.M.; Høyer, S.; Lynnerup, A.S.; Ørntoft, T.F.; Sørensen, K.D.; Borre, M.; Dyrskjøt, L. Expression Profiling of Prostate Cancer Tissue Delineates Genes Associated with Recurrence after Prostatectomy. Sci. Rep. 2015, 5, doi:10.1038/srep16018.

23. Wallace, T.A.; Prueitt, R.L.; Yi, M.; Howe, T.M.; Gillespie, J.W.; Yfantis, H.G.; Stephens, R.M.; Caporaso, N.E.; Loffredo, C.A.; Ambs, S. Tumor Immunobiological Differences in Prostate Cancer between African-American and European-American Men. Cancer Res. 2008, 68, 927–936, doi:10.1158/0008-5472.CAN-07-2608.

24. Ross-Adams, H.; Lamb, A.; Dunning, M.; Halim, S.; Lindberg, J.; Massie, C.; Egevad, L.; Russell, R.; Ramos-Montoya, A.; Vowler, S.;, et al. Integration of Copy Number and Transcriptomics Provides Risk Stratification in Prostate Cancer: A Discovery and Validation Cohort Study. EBioMedicine 2015, 2, 1133–1144, doi:10.1016/j.ebiom.2015.07.017.

25. TCGA-PRAD Available online: https://portal.gdc.cancer.gov/projects/TCGA-PRAD (accessed on 4 August 2021).

26. Taylor, B.S.; Schultz, N.; Hieronymus, H.; Gopalan, A.; Xiao, Y.; Carver, B.S.; Arora, V.K.; Kaushik, P.; Cerami, E.; Reva, B.;, et al. Integrative Genomic Profiling of Human Prostate Cancer. Cancer Cell 2010, 18, 11–22, doi:10.1016/j.ccr.2010.05.026.

27. Shi, W.; Oshlack, A.; Smyth, G.K. Optimizing the Noise versus Bias Trade-off for Illumina Whole Genome Expression BeadChips. Nucleic Acids Res. 2010, 38, e204, doi:10.1093/nar/gkq871.

28. Law, C.W.; Chen, Y.; Shi, W.; Smyth, G.K. Voom: Precision Weights Unlock Linear Model Analysis Tools for RNA-Seq Read Counts. Genome Biol. 2014, 15, R29, doi:10.1186/gb-2014-15-2-r29.

29. Jain, S.; Lyons, C.A.; Walker, S.M.; McQuaid, S.; Hynes, S.O.; Mitchell, D.M.; Pang, B.; Logan, G.E.; McCavigan, A.M.; O’Rourke, D.;, et al. Validation of a Metastatic Assay Using Biopsies to Improve Risk Stratification in Patients with Prostate Cancer Treated with Radical Radiation Therapy. Ann. Oncol. Off. J. Eur. Soc. Med. Oncol. 2018, 29, 215–222, doi:10.1093/annonc/mdx637.

30. Sboner, A.; Demichelis, F.; Calza, S.; Pawitan, Y.; Setlur, S.R.; Hoshida, Y.; Perner, S.; Adami, H.O.; Fall, K.; Mucci, L.A.;, et al. Molecular Sampling of Prostate Cancer: A Dilemma for Predicting Disease Progression. BMC Med. Genomics 2010, 3, doi:10.1186/1755-8794-3-8.

31. Long, Q.; Xu, J.; Osunkoya, A.O.; Sannigrahi, S.; Johnson, B.A.; Zhou, W.; Gillespie, T.; Park, J.Y.; Nam, R.K.; Sugar, L.;, et al. Global Transcriptome Analysis of Formalin-Fixed Prostate Cancer Specimens Identifies Biomarkers of Disease Recurrence. Cancer Res. 2014, 74, 3228–3237, doi:10.1158/0008-5472.CAN-13-2699.

32. Luca, B.-A.; Brewer, D.S.; Edwards, D.R.; Edwards, S.; Whitaker, H.C.; Merson, S.; Dennis, N.; Cooper, R.A.; Hazell, S.; Warren, A.Y.;, et al. DESNT: A Poor Prognosis Category of Human Prostate Cancer. Eur. Urol. Focus 2018, 4, 842–850, doi:10.1016/j.euf.2017.01.016.

33. Gerhauser, C.; Favero, F.; Risch, T.; Simon, R.; Feuerbach, L.; Assenov, Y.; Heckmann, D.; Sidiropoulos, N.; Waszak, S.M.; Hübschmann, D.;, et al. Molecular Evolution of Early-Onset Prostate Cancer Identifies Molecular Risk Markers and Clinical Trajectories. Cancer Cell 2018, 34, 996–1011.e8, doi:10.1016/j.ccell.2018.10.016.

34. Abida, W.; Cyrta, J.; Heller, G.; Prandi, D.; Armenia, J.; Coleman, I.; Cieslik, M.; Benelli, M.; Robinson, D.; Van Allen, E.M.;, et al. Genomic Correlates of Clinical Outcome in Advanced Prostate Cancer. Proc. Natl. Acad. Sci. U. S. A. 2019, 116, 11428–11436, doi:10.1073/pnas.1902651116.

35. Bland, J.M.; Altman, D.G. The Logrank Test. BMJ 2004, 328, 1073, doi:10.1136/bmj.328.7447.1073.

36. Budczies, J.; Klauschen, F.; Sinn, B.V.; Gyorffy, B.; Schmitt, W.D.; Darb-Esfahani, S.; Denkert, C. Cutoff Finder: A Comprehensive and Straightforward Web Application Enabling Rapid Biomarker Cutoff Optimization. PLoS ONE 2012, 7, 1–7, doi:10.1371/journal.pone.0051862.

37. Breslow, N.E. Analysis of Survival Data under the Proportional Hazards Model. Int. Stat. Rev. Rev. Int. Stat. 1975, 43, 45–57, doi:10.2307/1402659.

38. Therneau, T. A Package for Survival Analysis in S. R Package Version. Survival 2012.

39. Kassambara, A.; Kosinski, M.; Biecek, P.; Fabian, S. Package ‘Survminer’. Drawing Survival Curves Using ‘Ggplot2’. R Package Version 0.3.1, https://CRAN.R-Project.Org/Package=survminer. *null* 2014.

40. Ishwaran, H.; Kogalur, U.B. randomForestSRC: Fast Unified Random Forests for Survival, Regression, and Classification (RF-SRC) 2024.

41. Ishwaran, H.; Lu, M. Standard Errors and Confidence Intervals for Variable Importance in Random Forest Regression, Classification, and Survival. Stat. Med. 2019, 38, 558–582, doi:10.1002/sim.7803.

42. Palanisamy, N.; Yang, J.; Shepherd, P.D.A.; Li-Ning-Tapia, E.M.; Labanca, E.; Manyam, G.C.; Ravoori, M.K.; Kundra, V.; Araujo, J.C.; Efstathiou, E.;, et al. The MD Anderson Prostate Cancer Patient-Derived Xenograft Series (MDA PCa PDX) Captures the Molecular Landscape of Prostate Cancer and Facilitates Marker-Driven Therapy Development. Clin. Cancer Res. 2020, 26, 4933–4946, doi:10.1158/1078-0432.CCR-20-0479.

43. Anselmino, N.; Labanca, E.; Shepherd, P.D.A.; Dong, J.; Yang, J.; Song, X.; Nandakumar, S.; Kundra, R.; Lee, C.; Schultz, N.;, et al. Integrative Molecular Analyses of the MD Anderson Prostate Cancer Patient-Derived Xenograft (MDA PCa PDX) Series. Clin. Cancer Res. Off. J. Am. Assoc. Cancer Res. 2024, 30, 2272–2285, doi:10.1158/1078-0432.CCR-23-2438.

44. Kolde, R. Pheatmap: Pretty Heatmaps 2019.

45. Kassambara, A.; Mundt, F. Factoextra: Extract and Visualize the Results of Multivariate Data Analyses. R Package. 2020.

46. Robin, X.; Turck, N.; Hainard, A.; Tiberti, N.; Lisacek, F.; Sanchez, J.-C.; Müller, M. pROC: Display and Analyze ROC Curves 2010, 1.18.5.

47. Cerami, E.; Gao, J.; Dogrusoz, U.; Gross, B.E.; Sumer, S.O.; Aksoy, B.A.; Jacobsen, A.; Byrne, C.J.; Heuer, M.L.; Larsson, E.;, et al. The cBio Cancer Genomics Portal: An Open Platform for Exploring Multidimensional Cancer Genomics Data. Cancer Discov. 2012, 2, 401–404, doi:10.1158/2159-8290.CD-12-0095.

48. Gao, J.; Aksoy, B.A.; Dogrusoz, U.; Dresdner, G.; Gross, B.; Sumer, S.O.; Sun, Y.; Jacobsen, A.; Sinha, R.; Larsson, E.;, et al. Integrative Analysis of Complex Cancer Genomics and Clinical Profiles Using the cBioPortal. Sci. Signal. 2013, 6, doi:10.1126/scisignal.2004088.

49. de Bruijn, I.; Kundra, R.; Mastrogiacomo, B.; Tran, T.N.; Sikina, L.; Mazor, T.; Li, X.; Ochoa, A.; Zhao, G.; Lai, B.;, et al. Analysis and Visualization of Longitudinal Genomic and Clinical Data from the AACR Project GENIE Biopharma Collaborative in cBioPortal. Cancer Res. 2023, 83, 3861–3867, doi:10.1158/0008-5472.CAN-23-0816.

50. Dexter, T.A. R: A Language and Environment for Statistical Computing. Quat. Res. 2014, 81, 114–124, doi:10.1016/j.quascirev.2005.03.008.

51. RStudio RStudio | Open Source & Professional Software for Data Science Teams –RStudio Available online: https://www.rstudio.com/ (accessed on 22 September 2021).

52. Wickham, H.; Averick, M.; Bryan, J.; Chang, W.; McGowan, L.; François, R.; Grolemund, G.; Hayes, A.; Henry, L.; Hester, J.;, et al. Welcome to the Tidyverse. J. Open Source Softw. 2019, 4, 1686, doi:10.21105/joss.01686.

53. Wickham; Hadley Ggplot2. Elegant Graphics for Data Analysis; Springer-Verlag: NY, 2016; ISBN 978-3-319-24277-4.

54. Kassambara, A. Ggpubr: “ggplot2” Based Publication Ready Plots.R Package Version 0.4.0.999. https://Rpkgs.Datanovia.Com/Ggpubr/2020.

55. Neuwirth, E.; Maindonald, J. Package “RColorBrewer.” 2015.

56. Davis, S.; Meltzer, P.S. GEOquery: A Bridge between the Gene Expression Omnibus (GEO) and BioConductor. Bioinforma. Oxf. Engl. 2007, 23, 1846–1847, doi:10.1093/bioinformatics/btm254.

57. Serritella, A.V.; Beltran, H.; Lotan, T.L.; VanderWeele, D.J.; Karzai, F.; Madan, R.A.; Hussain, M. Large Cell Neuroendocrine Prostate Cancer: Large Is Not Small. The Oncologist 2024, 29, 185–189, doi:10.1093/oncolo/oyad344.

58. Epstein, J.I.; Amin, M.B.; Beltran, H.; Lotan, T.L.; Mosquera, J.-M.; Reuter, V.E.; Robinson, B.D.; Troncoso, P.; Rubin, M.A. Proposed Morphologic Classification of Prostate Cancer With Neuroendocrine Differentiation. Am. J. Surg. Pathol. 2014, 38, 756–767, doi:10.1097/PAS.0000000000000208.

59. Humphrey, P.A.; Moch, H.; Cubilla, A.L.; Ulbright, T.M.; Reuter, V.E. The 2016 WHO Classification of Tumours of the Urinary System and Male Genital Organs—Part B: Prostate and Bladder Tumours. Eur. Urol. 2016, 70, 106–119, doi:10.1016/j.eururo.2016.02.028.

60. Evans, A.J.; Humphrey, P.A.; Belani, J.; van der Kwast, T.H.; Srigley, J.R. Large Cell Neuroendocrine Carcinoma of Prostate: A Clinicopathologic Summary of 7 Cases of a Rare Manifestation of Advanced Prostate Cancer. Am. J. Surg. Pathol. 2006, 30, 684–693, doi:10.1097/00000478-200606000-00003.

61. Nguyen, N.; Ronald Dean Franz, I.I.; Mohammed, O.; Huynh, R.; Son, C.K.; Khan, R.N.; Ahmed, B. A Systematic Review of Primary Large Cell Neuroendocrine Carcinoma of the Prostate. Front. Oncol. 2024, 14, doi:10.3389/fonc.2024.1341794.

62. Aggarwal, R.; Zhang, T.; Small, E.J.; Armstrong, A.J. Neuroendocrine Prostate Cancer: Subtypes, Biology, and Clinical Outcomes. J. Natl. Compr. Cancer Netw. JNCCN 2014, 12, 719–726, doi:10.6004/jnccn.2014.0073.

63. Bhagirath, D.; Liston, M.; Akoto, T.; Lui, B.; Bensing, B.A.; Sharma, A.; Saini, S. Novel, Non-Invasive Markers for Detecting Therapy Induced Neuroendocrine Differentiation in Castration-Resistant Prostate Cancer Patients. Sci. Rep. 2021, 11, 8279, doi:10.1038/s41598-021-87441-2.

64. Dang, Q.; Li, L.; Xie, H.; He, D.; Chen, J.; Song, W.; Chang, L.S.; Chang, H.-C.; Yeh, S.; Chang, C. Anti-Androgen Enzalutamide Enhances Prostate Cancer Neuroendocrine (NE) Differentiation via Altering the Infiltrated Mast Cells → Androgen Receptor (AR) → miRNA32 Signals. Mol. Oncol. 2015, 9, 1241–1251, doi:10.1016/j.molonc.2015.02.010.

65. Ou, Y.-H.; Jiang, Y.-D.; Li, Q.; Zhuang, Y.-J.; Dang, Q.; Tan, W.-L. [Infiltrating mast cells promote neuroendocrine differentiation and increase docetaxel resistance of prostate cancer cells by up-regulating p21]. Nan Fang Yi Ke Da Xue Xue Bao 2018, 38, 723–730, doi:10.3969/j.issn.1673-4254.2018.06.13.

66. Maimaitiyiming, A.; An, H.; Xing, C.; Li, X.; Li, Z.; Bai, J.; Luo, C.; Zhuo, T.; Huang, X.; Maimaiti, A.;, et al. Machine Learning-Driven Mast Cell Gene Signatures for Prognostic and Therapeutic Prediction in Prostate Cancer. Heliyon 2024, 10, doi:10.1016/j.heliyon.2024.e35157.

67. Aller, M.-A.; Arias, A.; Arias, J.-I.; Arias, J. Carcinogenesis: The Cancer Cell–Mast Cell Connection. Inflamm. Res. 2019, 68, 103–116, doi:10.1007/s00011-018-1201-4.

68. Conteduca, V.; Oromendia, C.; Eng, K.W.; Bareja, R.; Sigouros, M.; Molina, A.; Faltas, B.M.; Sboner, A.; Mosquera, J.M.; Elemento, O.;, et al. Clinical Features of Neuroendocrine Prostate Cancer. Eur. J. Cancer 2019, 121, 7–18, doi:10.1016/j.ejca.2019.08.011.

69. Cherif, C.; Nguyen, D.T.; Paris, C.; Le, T.K.; Sefiane, T.; Carbuccia, N.; Finetti, P.; Chaffanet, M.; Kaoutari, A.E.; Vernerey, J.;, et al. Menin Inhibition Suppresses Castration-Resistant Prostate Cancer and Enhances Chemosensitivity. Oncogene 2022, 41, 125–137, doi:10.1038/s41388-021-02039-2.

70. Quan, Y.; Zhang, X.; Wang, M.; Ping, H. Histone Lysine Methylation Patterns in Prostate Cancer Microenvironment Infiltration: Integrated Bioinformatic Analysis and Histological Validation. Front. Oncol. 2022, 12, 981226, doi:10.3389/fonc.2022.981226.

71. Tzelepi, V.; Logotheti, S.; Efstathiou, E.; Troncoso, P.; Aparicio, A.; Sakellakis, M.; Hoang, A.; Perimenis, P.; Melachrinou, M.; Logothetis, C.;, et al. Epigenetics and Prostate Cancer: Defining the Timing of DNA Methyltransferase Deregulation during Prostate Cancer Progression. Pathology (Phila.) 2020, 52, 218–227, doi:10.1016/j.pathol.2019.10.006.

72. Enz, N.; Vliegen, G.; De Meester, I.; Jungraithmayr, W. CD26/DPP4 – a Potential Biomarker and Target for Cancer Therapy. Pharmacol. Ther. 2019, 198, 135–159, doi:10.1016/j.pharmthera.2019.02.015.

73. Burdelski, C.; Strauss, C.; Tsourlakis, M.C.; Kluth, M.; Hube-Magg, C.; Melling, N.; Lebok, P.; Minner, S.; Koop, C.; Graefen, M.;, et al. Overexpression of Thymidylate Synthase (TYMS) Is Associated with Aggressive Tumor Features and Early PSA Recurrence in Prostate Cancer. Oncotarget 2015, 6, 8377–8387, doi:10.18632/oncotarget.3107.

74. Ngan, E.S.W.; Hashimoto, Y.; Ma, Z.-Q.; Tsai, M.-J.; Tsai, S.Y. Overexpression of Cdc25B, an Androgen Receptor Coactivator, in Prostate Cancer. Oncogene 2003, 22, 734–739, doi:10.1038/sj.onc.1206121.

75. Roberts, B.K.; Collado, G.; Barnes, B.J. Role of Interferon Regulatory Factor 5 (IRF5) in Tumor Progression: Prognostic and Therapeutic Potential. Biochim. Biophys. Acta BBA – Rev. Cancer 2024, 1879, 189061, doi:10.1016/j.bbcan.2023.189061.

76. Chen, H.; Fang, S.; Zhu, X.; Liu, H. Cancer-Associated Fibroblasts and Prostate Cancer Stem Cells: Crosstalk Mechanisms and Implications for Disease Progression. Front. Cell Dev. Biol. 2024, 12, doi:10.3389/fcell.2024.1412337.

77. Ellis, L.; Loda, M. Advanced Neuroendocrine Prostate Tumors Regress to Stemness. Proc. Natl. Acad. Sci. 2015, 112, 14406–14407, doi:10.1073/pnas.1519151112.

78. Chakraborty, G.; Gupta, K.; Kyprianou, N. Epigenetic Mechanisms Underlying Subtype Heterogeneity and Tumor Recurrence in Prostate Cancer. Nat. Commun. 2023, 14, 567, doi:10.1038/s41467-023-36253-1.

79. Guijarro, M.V.; Nawab, A.; Dib, P.; Burkett, S.; Luo, X.; Feely, M.; Nasri, E.; Seifert, R.P.; Kaye, F.J.; Zajac-Kaye, M. TYMS Promotes Genomic Instability and Tumor Progression in Ink4a/Arf Null Background. Oncogene 2023, 42, 1926–1939, doi:10.1038/s41388-023-02694-7.

80. Ibe, T.; Shimizu, K.; Nakano, T.; Kakegawa, S.; Kamiyoshihara, M.; Nakajima, T.; Kaira, K.; Takeyoshi, I. High-Grade Neuroendocrine Carcinoma of the Lung Shows Increased Thymidylate Synthase Expression Compared to Other Histotypes. J. Surg. Oncol. 2010, 102, 11–17, doi:10.1002/jso.21576.

81. Gao, H.; Korn, J.M.; Ferretti, S.; Monahan, J.E.; Wang, Y.; Singh, M.; Zhang, C.; Schnell, C.; Yang, G.; Zhang, Y.;, et al. High-Throughput Screening Using Patient-Derived Tumor Xenografts to Predict Clinical Trial Drug Response. Nat. Med. 2015, 21, 1318–1325, doi:10.1038/nm.3954.

